# External validation, recalibration and updating of the OxSATS risk model for suicide after self-harm in England

**DOI:** 10.64898/2026.01.28.26345038

**Authors:** Tyra Lagerberg, Denis Yukhnenko, Maria Vazquez-Montes, Thomas R. Fanshawe, Seena Fazel

## Abstract

**Background:** External validations of existing risk models is an efficient step towards potential implementation, obviating the need to develop new models. However, validation in new clinical settings poses several challenges.

**Objective:** To externally validate the OxSATS tool using data from the Oxford Monitoring System for Self-harm in England. OxSATS is a validated tool to predict suicide after self-harm developed using Swedish population registers.

**Methods:** We selected episodes of self-harm (ICD-10 codes X60-84; Y10-34) by individuals aged 10-64 years who presented to a large regional hospital between 1 January 2000 and 31 December 2018, and were followed up until 31 December 2019. We applied the OxSATS tool to estimate each individual’s suicide risk within 12 months after their index self-harm. We assessed model performance using discrimination (Harrell’s c-index) and calibration measures (calibration plot and the observed-to-expected events ratio, O:E). We assessed the effects of missing predictors on calibration and subsequently recalibrated the model.

**Findings:** We identified 16,120 individuals who presented to hospital with self-harm, of whom 101 (0.6%) died by suicide in the 12-month follow-up period. The OxSATS model showed good discrimination in external validation (c-index=0.72, 95% CI=0.67, 0.77). Recalibration was required because initial calibration reflected a lower outcome rate in the new data. After recalibration, calibration performance was excellent (O:E=1.00, 95% CI=0.80, 1.20).

**Conclusions:** Despite differences in clinical services and outcome ascertainment, suicide risk models can maintain good predictive performance in new settings. However, recalibration should be considered when applying prediction models in new settings, and the impact of missing predictors should be assessed using sensitivity analyses.

**KEY MESSAGES:** *What is already known on this topic:* Suicide risk is substantially elevated after hospital presentation for self-harm, but most existing risk assessment tools rely on rating scales or binary cut-offs, show limited predictive accuracy, and rarely report calibration. OxSATS is a prognostic model developed using Swedish register data that provides continuous risk estimates and demonstrated good discrimination and calibration in its original setting. External validation in new healthcare systems is essential before implementation, but is often complicated by differences in predictor definitions, missing variables, and outcome prevalence.

*What this study adds:* This study provides the first external validation of OxSATS in an English clinical setting using routinely collected hospital data. The model retained good discrimination but initially overpredicted suicide risk due to a lower baseline event rate and one missing predictor, highlighting the importance of calibration assessment.

*How this study might affect research, practice or policy:* Future research and implementation strategies should routinely incorporate external validation, sensitivity analyses for missing predictors, and local recalibration before clinical or policy adoption.

## INTRODUCTION

Suicide is a leading cause of death, particularly among children and young adults (Fazel & Runeson, 2020). A major risk marker for suicide is prior self-harm – between 1% and 2% of those who present to clinical services with self-harm go on to die by suicide over the next 12 months (Fazel et al., 2023; Olfson et al., 2017). Identifying individuals at particularly elevated risk in this population can potentially inform decision-making around risk management, resource allocation, and follow-up.

The Oxford Suicide Assessment Tool for Self-harm (OxSATS) was developed using Swedish national population registers to estimate suicide risk after presentation with self-harm to emergency departments and secondary mental health services. The risk model showed good predictive performance in Swedish validation in relation to discrimination (c-index=0.77, 95%CI=0.75-0.78) and calibration (O:E=0.92, 95% CI=0.79, 1.06) and provides a continuous risk score for flexible application in research and clinical care. The output of continuous probability scores is a key difference to previous tools, which have relied on binary cut-offs and evaluation of performance only on classification measures (such as sensitivity and specificity) while neglecting to consider model calibration and overall discrimination. Another difference is that almost all previous tools used in practice are rating scales not developed or intended for prognostic probability prediction (Fazel et al., 2023). OxSATS, in contrast, was developed using high quality methods for prediction modelling, including a pre-specified analytic protocol, empirically derived predictors, clear prediction horizons, and a large population dataset, ensuring outcome numbers sufficient for model development. Predictors were chosen to be generalisable to different settings. Reporting of the model was comprehensive and transparent, following established guidelines.

Nevertheless, implementation of OxSATS for research, training and clinical purposes in settings outside Sweden should test its predictive performance in the new intended population (external validation) beforehand and recalibrate the model if poor calibration is demonstrated. In a new country or setting, using a tool already developed is an efficient research approach, as it does not require investment in developing a new tool, which requires larger data sets (Steyerberg, 2009). We therefore aimed to test the performance of the OxSATS risk model using data from a regional database in England, the Oxford Monitoring System for Self-harm. Validating prediction models in new settings is associated with challenges, including different predictor definitions, missing predictors, and a base event rate and outcome definition that can vary from the original sample used for model development.

## METHODS

### Data sources

The Oxford Monitoring System for Self-harm contains data of individuals who presented to an emergency department with a non-fatal self-harm event at a large general hospital in Oxford (Centre for Suicide Research, 2025).

### Study population

We included individuals aged 10 or above who presented to an emergency department with an episode of self-harm between 1 January 2000 and 31 December 2018. In individuals with more than one self-harm episode, we randomly selected a single episode as the index, in keeping with previous work and the original model development (Fazel et al., 2023). We followed up individuals for 12 months up to 31 December 2019, from the date of their index self-harm episode to determine whether they died by suicide during that period.

### Prediction model

The OxSATS risk model was developed and validated in 53,172 individuals aged at least 10 who presented to specialist services for non-fatal self-harm in Sweden between 2008 and 2012 (Fazel et al., 2023). It includes 11 routinely collected predictors and is available as an online risk calculator at https://oxrisk.com/oxsats/.

### Definition of outcomes

The outcome was death by suicide (ICD-10 codes X60-X84 and Y10-Y34) within 12 months of presentation with non-fatal self-harm to specialist services. Y10-Y34 codes (deaths from external causes of unknown intent) were included in the outcome to avoid undercounting and misclassification (Fazel et al., 2023).

### Definition of predictors

All predictors in OxSATS were present or had an equivalent in the Oxford Monitoring System for Self-harm data apart from “any psychotropic medication receipt [collection of a prescription] in past 3 months” (current psychotropic medication use). The predictors included age, sex, current or lifetime alcohol use disorder, current of lifetime drug use disorder, prior self-harm (psychotropic drug overdose; any prior hanging strangulation, or suffocation; any prior self-harm within 12 months of the index episode), any psychiatric disorder except for substance use disorder. The covariates were collected at the time of admission by clinicians, were assumed to be complete, and no missing data imputation was performed.

One predictor from the original model was not available, which was current use of any psychotropic medication. We accounted for the impact of this missing predictor through sensitivity analyses assuming different prevalence levels in the population (see Statistical Methods). We compared prevalence of predictors available in the OxSATS development sample and Oxford Monitoring data (see Results). One difference in definitions was that the OxSATS predictors for psychiatric diagnoses were based on lifetime diagnoses, while the equivalent predictors in Oxford Monitoring data were coded as mental health conditions that healthcare staff judged as contributing to the individual’s presenting act of self-harm.

### Power analysis

To determine the required sample size, we used the *pmvalsampsize* package for R, following the methodology for precision-based power analysis for external validation studies (Riley et al., 2024). Assuming an outcome prevalence of 0.6% and anticipated discrimination consistent with OxSATS, a sample size of 16,113 individuals (approximately 97 outcome events) would be expected to provide the following precision in performance estimates: overall calibration (O/E=1.00, 95% CI=0.80, 1.20), a calibration slope of 1.00 (95% CI=0.80, 1.20), and discrimination of c-index=0.77 (95% CI=0.70, 0.85). These calculations were used to characterise the expected precision of the validation analyses rather than to define a preregistered or fixed sample size target.

### Statistical methods

We tested predictive performance by estimating measures of discrimination and calibration. Discrimination measures included Harrell’s c-index, and the area under the receiver operating characteristic curve (AUC). Calibration was assessed using calibration plots and the ratio of overall observed to expected events (O:E ratio). As recommended by new TRIPOD-AI guidelines (Collins et al., 2024), we have not set predetermined thresholds for calibration and reported continuous calibration metrics. Calibration plots were created by plotting the mean predicted risk in 20 quantiles of the predicted probability against the proportion of observed events in each quantile (Fazel et al., 2024).

For the predictor missing (current psychotropic medication use), we set its value as equal to the prevalence of the predictor in the original OxSATS development sample, 0.557. As a sensitivity analysis, we tested the impact of randomly assigning the missing predictor as 0 or 1 in different individuals by using a Bernoulli distribution with p=0.557. We took the average of performance measures over 1,000 Monte Carlo simulations. In further sensitivity analyses, we assessed model performance by setting the missing predictor to values from 0 to 1 in steps of 0.1, to estimate how changes in the assumed probability influence calibration.

We initially evaluated the performance of OxSATS without updating the formula for calculating predicted risk. If calibration was found to be poor, we planned to update the intercept, shape parameter, and the predictors’ coefficients using a single multiplicative recalibration parameter. This procedure adjusts the calibration of the model while leaving the discrimination performance unchanged (Su et al., 2018). We did not plan to re-estimate individual predictor coefficients as that would require creating a new prediction tool.

All analyses were carried out using Python version 3.9.11. We followed TRIPOD-AI guidelines on the reporting of validation studies (for the checklist, refer to Supplemental).

### Ethical approval and data availability

The Oxford Self-Harm Monitoring System is compliant with the Data Protection Act 1998 and has approval under Section 251 of the National Health Service (NHS) Act 2006 to collect patient-identifiable information without explicit patient consent. The study rationale was discussed with the Centre for Suicide Research Public and Patient Involvement group. A separate study protocol was not published. The data are not publicly available and can be accessed only by researchers affiliated with the Oxford Self-Harm Monitoring System, subject to relevant governance approvals. The analysis code is available from the authors upon reasonable request.

## RESULTS

We identified 16,120 individuals who presented to an emergency department in a regional hospital centre with self-harm (Table 1). The mean age at the index self-harm episode was 31.5 years (SD 15.2), and 59.7% of individuals presenting with self-harm were female. A lifetime history of self-harm before the index episode was present in 35.6% persons, with 18.4% individuals having self-harmed in the 12 months preceding the index self-harm episode. Recorded mental health problems in the preceding 12 months were present in 12.2%. At index presentation, 26.5% involved an overdose with psychotropic medication, and 1.6% involved hanging, strangulation, or suffocation.

**Table 1.**
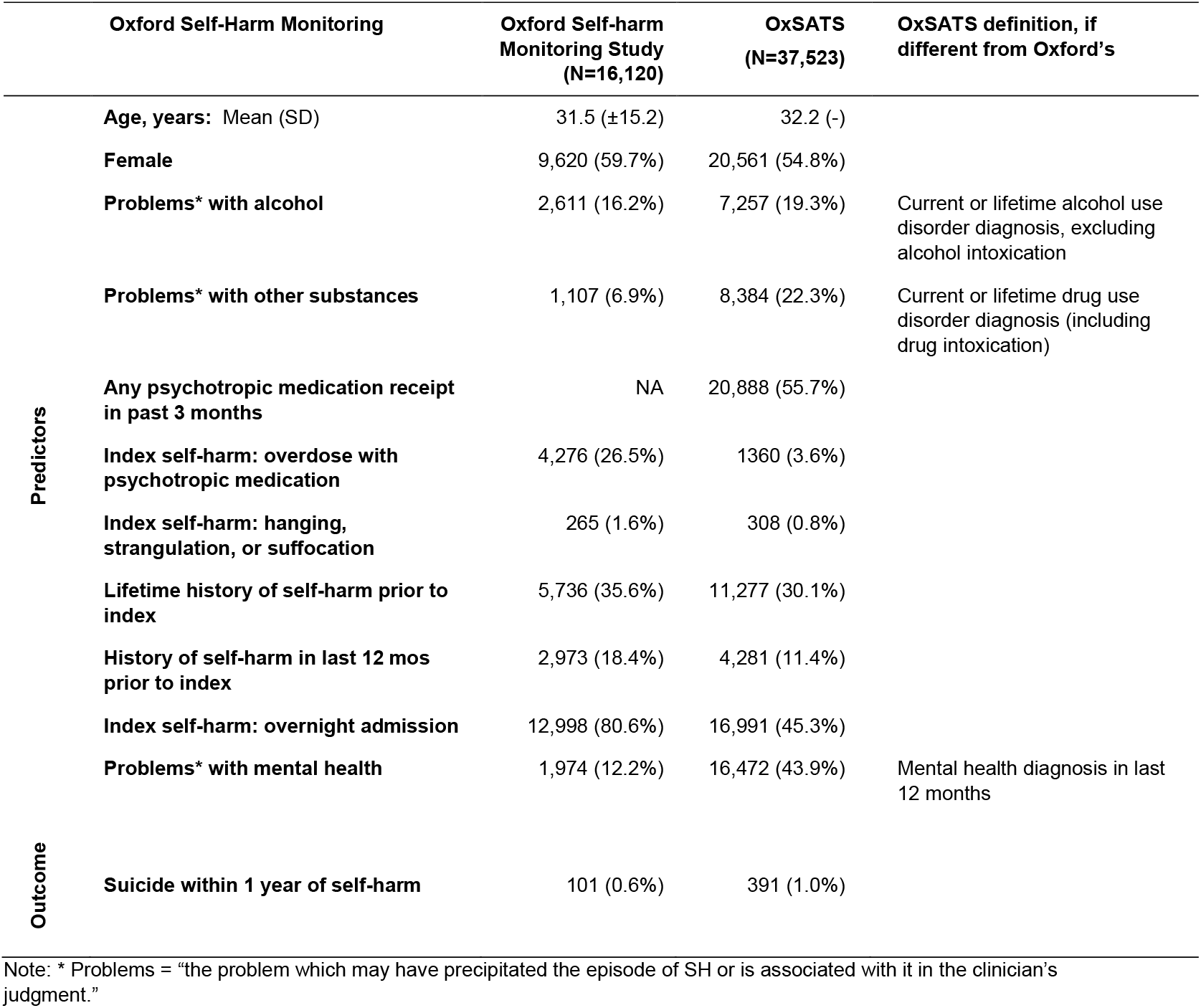
Prevalence of OxSATS predictors in the Oxford Monitoring System for Self-harm dataset, overall and stratified by whether individuals commit suicide within 1 year.

In Oxford Monitoring data, the outcome prevalence of death by suicide within 12 months of the index self-harm event was 0.6% (101/16120), lower than in the OxSATS development sample (1.0%; Table 1). Several predictors also showed differences in prevalence between the samples. These included any psychiatric diagnosis (12.2% in Oxford Monitoring data vs 43.9% in original OxSATS sample); problems with substances/substance misuse (6.9% vs 22.3%); and a history of self-harm (18.4% vs 11.4%).

Those who died by suicide within 12 months had a higher prevalence of all the predictors except problems with other substances; they were more likely to be male, have a prior history of self-harm, and method of self-harm as part of their current presentation (Table S1).

The discrimination performance of the OxSATS tool was good (c-index=0.72, 95% CI=0.67-0.77; AUC=0.72, 95% CI=0.67-0.72) (Table 2, Figure 1). However, the uncalibrated OxSATS model overestimated the risk across predicted probabilities, resulting in poor calibration (O:E=0.64, 95% CI=0.51, 0.76), with miscalibration being worse at higher probabilities (Figure S1).

**Table 2.** Discrimination performance in English compared to Swedish data after recalibration.

|  | Oxford self-harm data | OxSATS external validation sample |
| --- | --- | --- |
| Suicide within 1 year prevalence (95% CI) | 0.6% (0.5-0.8%) | 1.1% (1.0-1.3%) |
| C-index (95% CI) | 0.72 (0.67-0.77) | 0.77 (0.75-0.78) |
| AUC (95% CI) | 0.72 (0.67-0.77) | 0.74 (0.71-0.77) |

**Figure 1.**
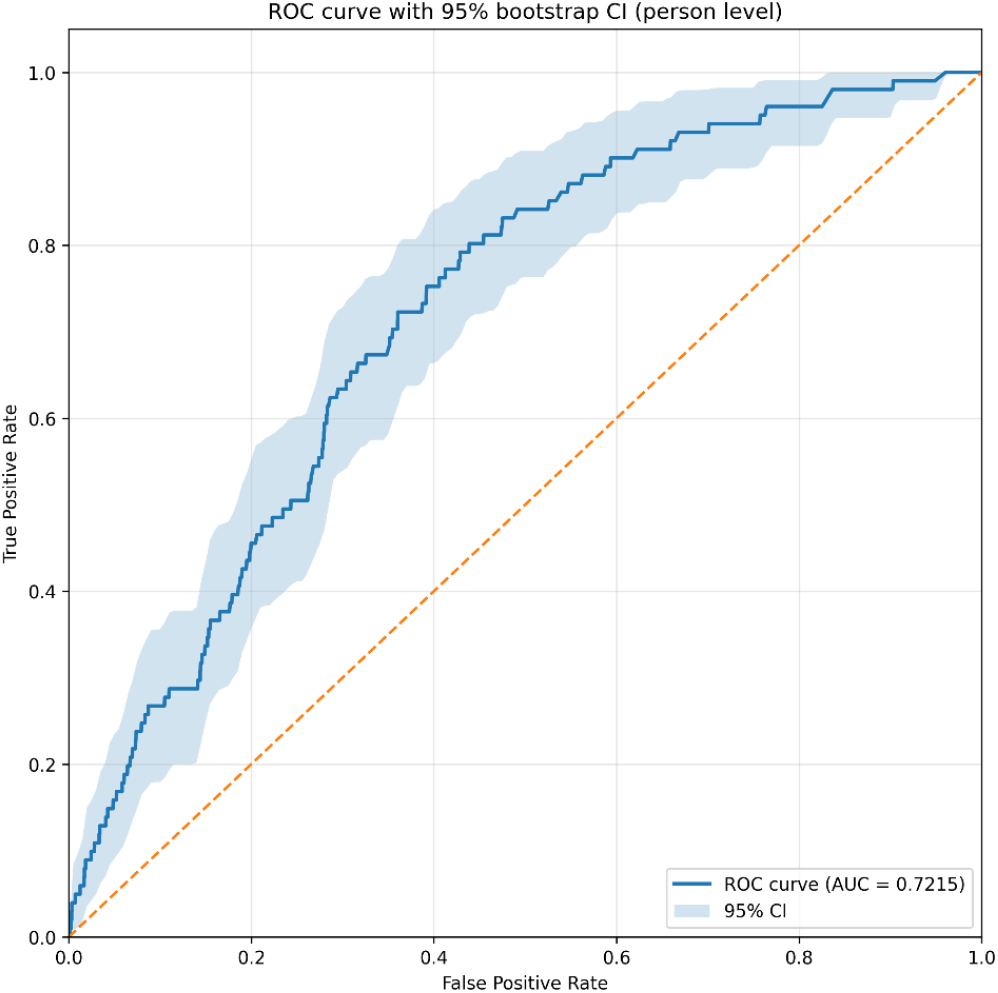
Receiver Operator Characteristics Curve of the OxSATS model in the Oxford Monitoring sample.

The missing predictor (current psychotropic medication use) was set to 0.557 in initial model assessment to reflect the prevalence in the OxSATS development sample. When we ran the Bernoulli simulation assigning 0 or 1 to each individual with p=0.557, the mean performance metrics over 1,000 simulations were similar to the main analysis, with a mean O:E of 0.59 (95% CI=0.58, 0.59) from the simulations, compared to 0.64 (95% CI=0.51, 0.76) in the main analysis (Table S2). The O:E ratio decreased monotonically as the assumed prevalence of the missing predictor increased (Figure S2), indicating increasing overprediction at higher prevalence levels.

We recalibrated the model by updating the intercept, shape parameter, and applying a fixed multiplier for the linear predictor (Table S3). As expected, this resulted in a well-calibrated model (O:E=1.00, 95% CI=0.80, 1.20; Figure 2).

**Figure 2.**
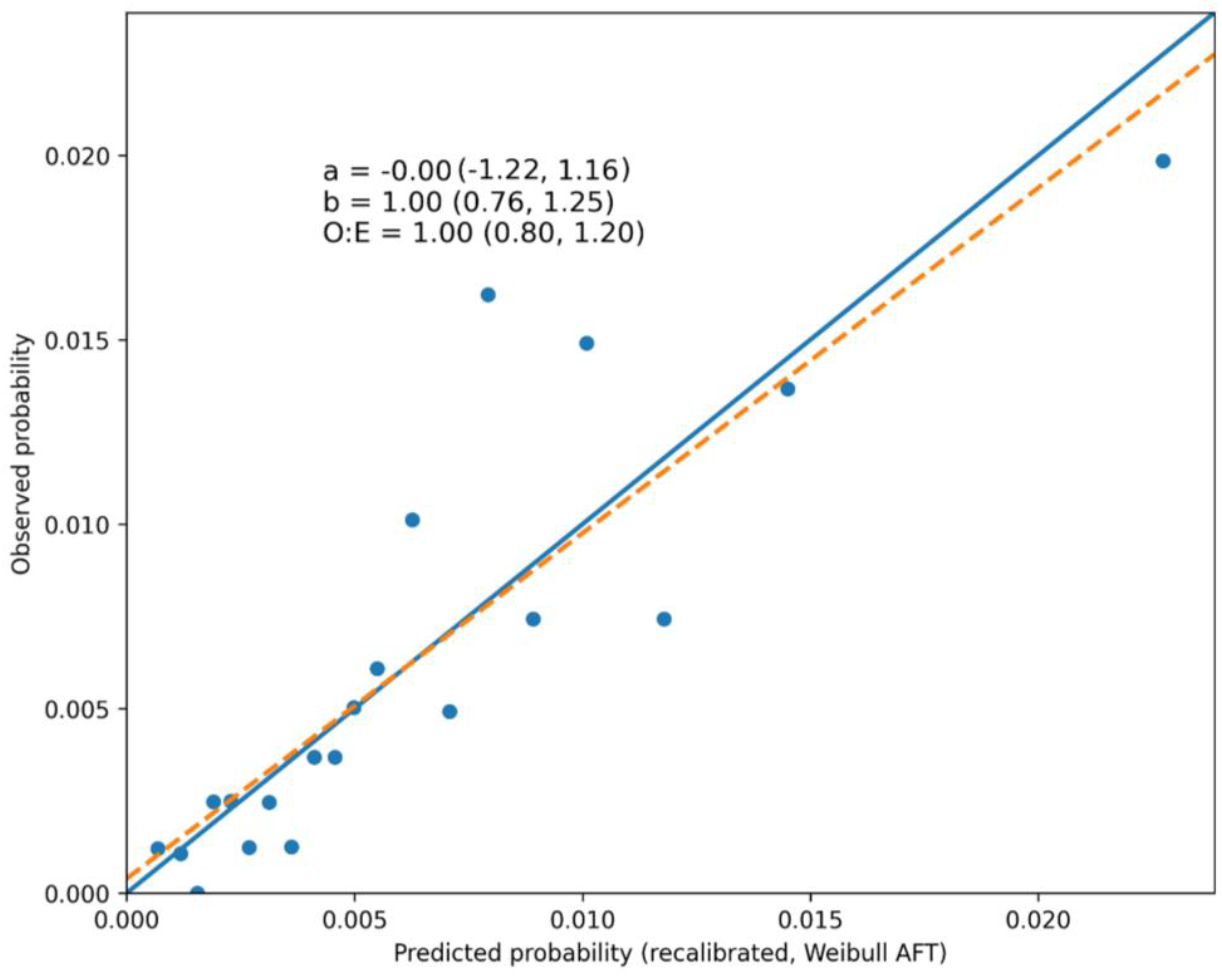
Calibration of OxSATS model in Oxford Monitoring data after recalibration.

## DISCUSSION

We investigated suicide mortality in 16,120 individuals who presented with a self-harm episode to a large regional English hospital. Over the follow-up of 12 months, 101 people died by suicide (0.6%). We tested the OxSATS tool to model suicide risk in the 12 months after the presenting self-harm episode and evaluated predictive performance using discrimination and calibration metrics. We found good overall discrimination performance (c-index=0.72, 95% CI=0.67, 0.77). However, there was a lower suicide rate after self-harm in this study than in the Swedish development sample, which meant calibration was initially poor, with the OxSATS tool over-predicting suicide risk (O:E=0.64). Recalibration successfully aligned the expected event rate with the average observed risk in this cohort (O:E=1.00, 95% CI=0.80, 1.20).

Estimating calibration thoroughly and considering recalibration is often overlooked as part of risk model validation (Seyedsalehi & Fazel, 2024). Unlike discrimination, which reflects only relative risk ranking, calibration determines whether absolute risk estimates generated by a risk model corresponds to real-world event rates and allows for interpretation of model outputs and potential actionability. Poorly calibrated models can systematically overpredict or underpredict risk. Overprediction can lead to unnecessarily restrictive interventions such as over-monitoring or hospitalization, while underprediction may result in insufficient follow-up and missed preventative opportunities. Ensuring good calibration is therefore essential for the safe and proportionate use of prediction models in clinical practice, especially for low-base-rate outcomes such as suicide.

### External validation and comparisons

Developing a new prediction tool for every new national context is costly and time-consuming – external validation of an existing tool offers an efficient use of resources. Additionally, there is often insufficient data to develop a prediction tool in new settings, particularly when considering rare outcomes. While we had adequate power over the 12-month horizon, there was insufficient power to externally validate the OxSATS tool at the 6-month time horizon.

The results of this external validation show some shrinkage in performance compared to the original Swedish study (c-index of 0.72 vs. 0.77), which would be expected given differences in national setting (England vs Sweden), data collection (questionnaires filled by healthcare staff vs linked national registers using routinely collected data) and one missing predictor. The good performance in external validation is in keeping with the OxSATS development: a simple, transparent model was preferred, and predictors were defined in ways expected to generalize to different national settings. Other tools developed using this overall approach have shown similarly strong discrimination performance in external validation. For example, the OxMIS tool, developed with Swedish data to model suicide risk in individuals with severe mental health problems (Fazel, Wolf, Larsson, et al., 2019), had an AUC of 0.70 (95% CI=0.69, 0.71) in an external validation using Finnish data (Sariaslan et al., 2023) as compared to 0.71 (95% CI=0.66, 0.75) in Swedish data. A recent review found that suicide risk prediction models show variable reduction in predictive performance when moving from internal to external validation, ranging from approximately 1% to 25% loss depending on the model used (derived from Seyedsalehi et al., 2025).

Despite these differences, OxSATS remains distinctive in the UK context as a model specifically developed and externally validated for suicide outcomes following self-harm, rather than for composite outcomes. In contrast, most existing tools have focused on combined outcomes of self-harm and suicide and have not reported calibration (Seyedsalehi et al., 2025).

In general, there are few studies that directly compare structured risk assessment tools with clinical judgment for the prediction of suicide and self-harm outcomes (Simon et al., 2021). In the largest study to date comparing clinician judgment with structured assessments, clinician prediction of suicide death was not demonstrably above chance, with an AUC=0.57 (95% CI =0.36, 0.73) (Randall et al., 2019). Risk models tools could therefore augment clinical judgment, supporting transparency and reproducibility in clinical assessment, and raise the ceiling of expertise by anchoring assessment in empirically derived models based on combining multiple risk markers. This is particularly relevant for services where access to trained mental health specialists is limited. In such settings, structured risk tools may be particularly useful by providing a consistent framework to support risk assessment alongside routine clinical care.

Examination of hybrid decision-making frameworks that formally combine probabilistic prediction with clinical judgment should be a priority for future research. One next step for research is to evaluate clinician accuracy both in isolation and when using statistical risk estimates (Simon et al., 2021). Such designs would allow separation of baseline clinical judgment, the incremental value added by risk models, and potential interaction effects, such as whether structured risk estimates improve calibration, consistency, or discrimination in clinicians’ assessments. This approach moves beyond simplistic dichotomies between “tools versus clinicians” and treats prediction as a sequential and collaborative process that more closely reflects real-world clinical workflows.

Using risk tools may also allow for more efficient resource allocation by making more explicit how various factors contribute to risk. There is an important difference, however, between the performance of a risk model, which is concerned with accuracy, and the decision about resource allocation, which will need to consider a range of individual and wider ethical and service-related factors. A health economic analysis has found that the OxMIS tool was cost-saving when integrated into clinician assessment of suicide risk in an English setting (Botchway et al., 2023) – similar assessments of the OxSATS model are required. In the US, studies find that positive predictive values lower than 1% for self-harm and less than 0.1% were found associated with cost effectiveness (Ross et al., 2021). To our knowledge, the accuracy of unaided clinician prediction of suicide following self-harm has not been investigated in the UK. Further work is called for in this area.

### Challenges of external validation

Our study demonstrates several challenges in externally validating a risk model in a new country. A first challenge is that some predictor definitions differed. In the current investigation, mental health variables were defined as problems that clinicians judged as directly related to the presenting self-harm, while in the OxSATS development sample they were defined as diagnosed lifetime psychiatric disorders prior to the index self-harm. This led to a lower prevalence of these variables in the Oxford Monitoring System for Self-harm data, and their effect was less strong but in a similar direction in the final multivariable model. Despite this, discriminative performance was good, and the reliance on proxy predictors meant that the reported performance is likely to present a conservative estimate of how well OxSATS performs (Fazel, Wolf, Vazquez-Montes, et al., 2019).

Another challenge was that one predictor was missing in the English data – current psychotropic medication use – which was an independent predictor in the original OxSATS model. Sensitivity analyses showed that assuming different prevalences for this predictor affected calibration performance. We assessed this by systematically varying the assumed prevalence and estimating calibration using bootstrap resampling, which allowed uncertainty arising from both sampling variability and assumptions about the missing predictor to be reflected in the performance estimates.

In addition to the effect of this missing predictor, differences in the prevalence of other predictors between datasets, such as medication overdose and overnight admission, may also have contributed to between-cohort differences in predicted risk and model performance. We found that there was a lower outcome prevalence in the English data compared to the Swedish development sample (0.6% vs 1.0% in Swedish data). This provides an explanation for why the OxSATS model initially over-predicted the risk of suicide after self-harm. Since recalibration lowered the predicted probabilities, the original OxSATS binary decision cut-offs are no longer directly applicable and would require local re-evaluation by the clinician or service that would be applying the tool.

## Conclusions

We have externally validated the OxSATS model in an English clinical setting and outlined a range of methodological challenges in validating risk models in new settings. These include approaches for handling a missing predictor and demonstrated the importance of model recalibration. Implementation strategies for prediction models should include external validation with transparent reporting of discrimination and calibration.

## Data Availability

The data are not publicly available and can be accessed only by researchers affiliated with the Oxford Self-Harm Monitoring System, subject to relevant governance approvals. The analysis code is available from the authors upon reasonable request.

