## Supplemental Material for "External validation, recalibration and updating of the OxSATS risk model for suicide after self-harm in England"

**SUPPLEMENT**

Table S1. Prevalence of OxSATS predictors in the Oxford Monitoring System for Self-harm dataset, overall and stratified by whether individuals commit suicide within 1 year

| | Participants without suicide within 1 year (N=16019) | Participants with suicide within 1 year (N=101) | All participants (N=16120) |
|---|---|---|---|
| **Age, years: Mean (SD)** | 31.5 (±15.1) | 42.6 (±16.9) | 31.5 (±15.2) |
| **Female** | 9588 (59.9%) | 32 (31.7%) | 9620 (59.7%) |
| **Problems* with alcohol** | 2592 (16.2%) | 19 (18.8%) | 2611 (16.2%) |
| **Problems* with other substances** | 1104 (6.9%) | 3 (3.0%) | 1107 (6.9%) |
| **Index self-harm: overdose with psychotropic medication** | 4241 (26.5%) | 35 (34.7%) | 4276 (26.5%) |
| **Index self-harm: hanging, strangulation, or suffocation** | 262 (1.6%) | 3 (3.0%) | 265 (1.6%) |
| **Lifetime history of self-harm prior to index** | 5692 (35.5%) | 44 (43.6%) | 5736 (35.6%) |
| **History of self-harm in last 12 mos prior to index** | 2943 (18.4%) | 30 (29.7%) | 2973 (18.4%) |
| **Index self-harm: overnight admission** | 12912 (80.6%) | 86 (85.1%) | 12998 (80.6%) |
| **Problems* with mental health** | 1954 (12.2%) | 20 (19.8%) | 1974 (12.2%) |

Note: * Problems = “the problem which may have precipitated the episode of SH or is associated with it in the clinician’s judgment.”

Table S2. Mean calibration metrics when randomly assigning the missing predictor as 1 or 0 to individuals in the study population following a Bernoulli distribution with p=0.557, compared to the main analysis

| | **Main analysis** | **Mean of 1000 simulations** |
|---|---|---|
| **O:E (95% CI)** | 0.63 (0.51, 0.75) | 0.59 (0.58, 0.59) |
| **Intercept** | 0.00 (0.00, 0.00) | 0.00 (0.00, 0.00) |
| **Slope** | 0.57 (0.37, 0.79) | 0.45 (0.36, 0.53) |

### Table S3. OxSATS model parameters and formula for probability of dying by suicide after self-harm presentation

| | | Parameter (β) | 95% confidence interval | |
|---|---|---|---|---|
| | **General demographics** | | | |
| | (Age at index/10)$^2$ | -4.20 | -5.32 | -3.08 |
| | Sex, female | -0.70 | -0.87 | -0.53 |
| | **Substance misuse** | | | |
| | Current or lifetime alcohol use disorder (excluding alcohol intoxication) | -0.03 | -0.22 | 0.17 |
| | Current or lifetime drug use disorder (including drug intoxication) | 0.31 | 0.12 | 0.50 |
| | **Treatment in the past three months** | | | |
| | Any psychotropic medication | 0.76 | 0.52 | 1.01 |
| | **History of self-harm** | | | |
| | Any psychotropic medication overdose | 0.42 | 0.02 | 0.82 |
| | Hanging, strangulation, or suffocation | 0.97 | 0.35 | 1.59 |
| | History of self-harm in the last 12 months prior to index, 1+ vs none | 0.31 | 0.07 | 0.55 |
| | Overnight admission | 0.57 | 0.38 | 0.75 |
| | **Mental health in the past 12 months** | | | |
| | Any psychiatric disorder except substance use disorders | 0.51 | 0.31 | 0.70 |
| | **Other parameters in the model – original model** | | | |
| | Intercept | -6.49 | -6.79 | -6.20 |
| | Shape parameter | 0.60 | 0.55 | 0.65 |
| | **Other parameters in the model – recalibrated model** | | | |
| **Recalibrated model** | Intercept | 4.37 | 1.77 | 6.98 |
| | Multiplier for linear predictor (original intercept + sum of risk factors multiplied by their parameters) | 1.70 | 1.15 | 2.26 |
| | Shape parameter | 0.57 | 0.47 | 0.70 |

Binary risk factors are coded as 1 if present and 0 if absent

In the original OxSATS tool, the risk for death by suicide within *t* months of presentation to specialist service with self-harm is calculated by the following formula:

$$\Pr(suicide\ within\ t\ months) = 1 - \exp\left(-\exp(linpred)\ t^{0.60}\right)$$

where $linpred = -6.49 + \sum \beta * Risk\ Factor$ and $t$ = 6 or 12

In the recalibrated tool, the risk for death by suicide within *t* months of self-harm is given by:

$$\Pr(suicide\ within\ t\ months) = 1 - \exp\left(-\exp(4.37 + 1.70 \times linpred)\ t^{0.57}\right)$$

where $linpred = -6.49 + \sum \beta * Risk\ Factor$ and $t$ = 12

In the recalibrated tool, the baseline survivor function is given by

$$S_0 = \exp\left(-\exp(-6.66) * t^{0.57}\right)$$

At 12 months this is $S_0(12) \approx 0.995$

Figure S1. Calibration of the original OxSATS tool in the Oxford Monitoring System for Self-harm data

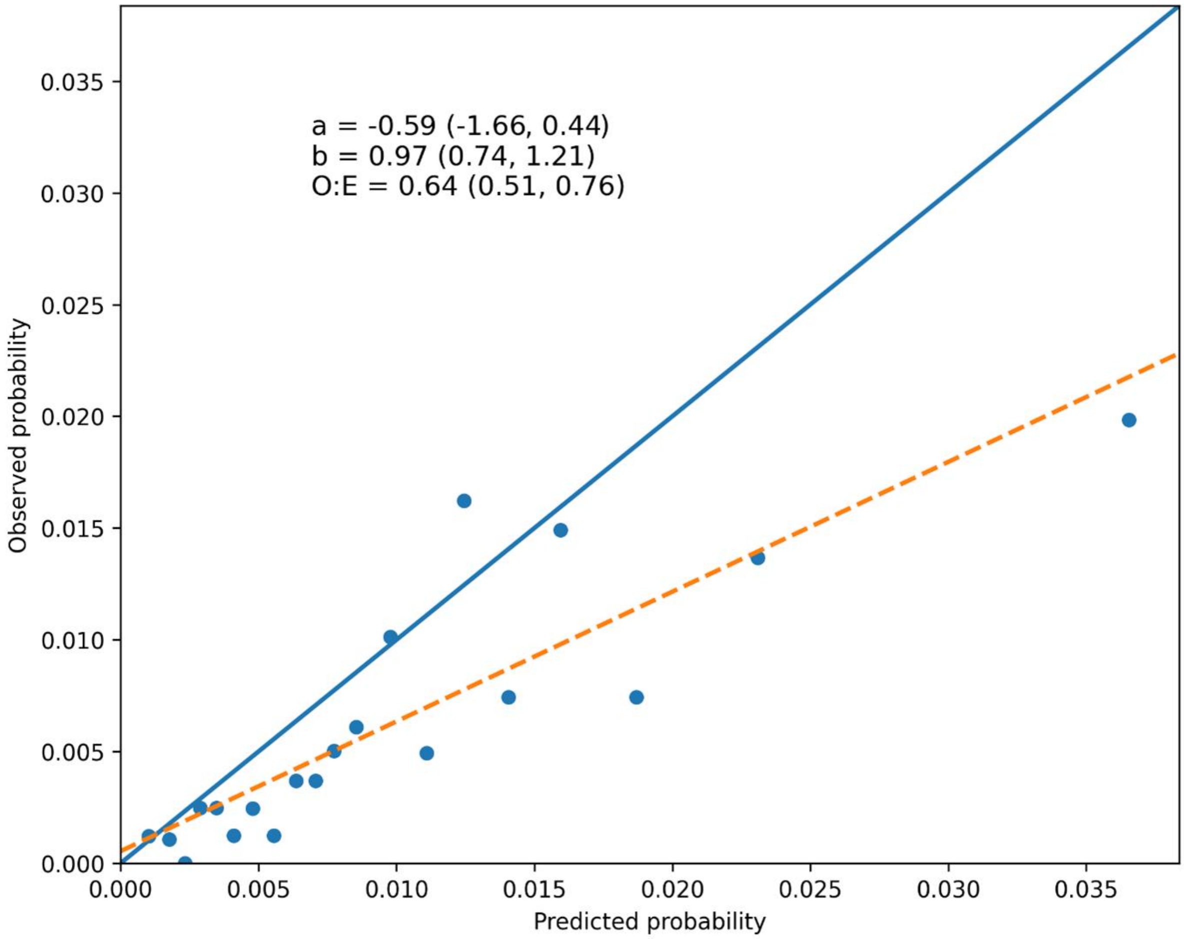

Figure S2. Calibration of OxSATS model in the Oxford Monitoring data when receipt of any psychotropic medication is changing from 0 to 1.

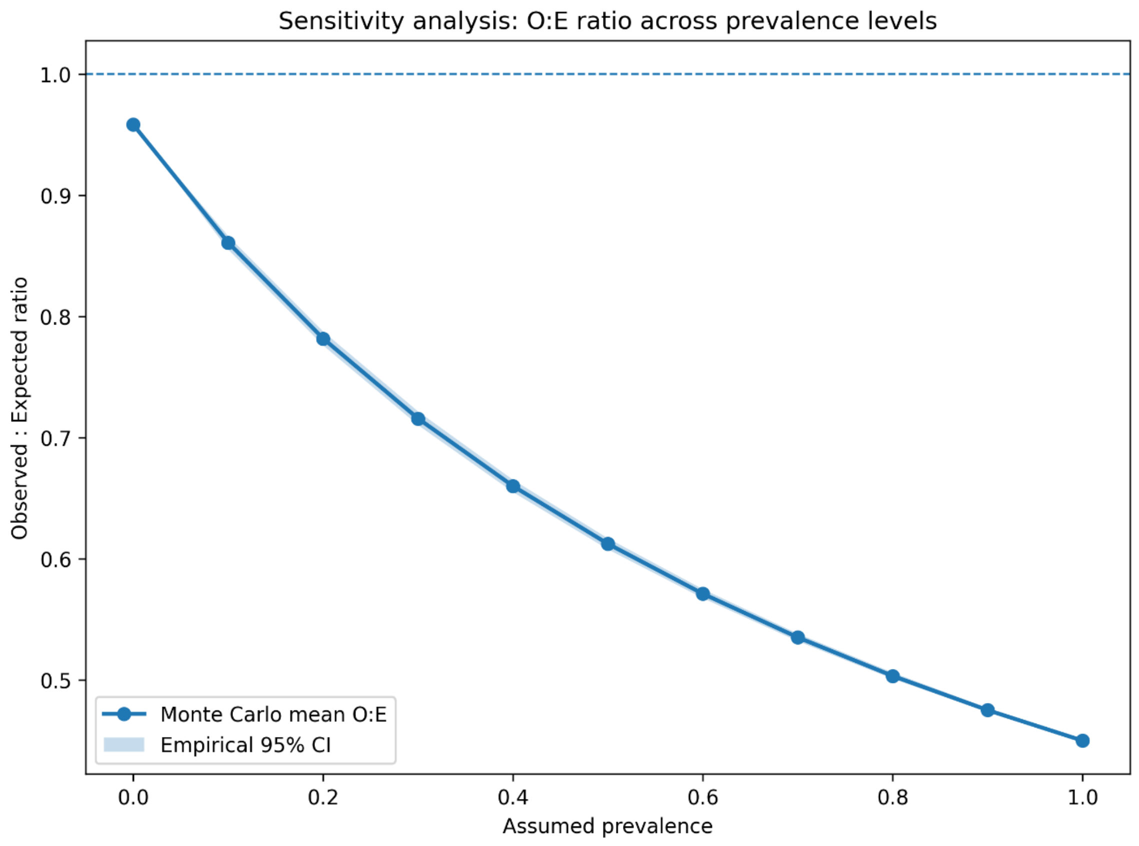
